# Cell type proportions rather than DNA methylation in the cord blood show significant associations with severe preeclampsia

**DOI:** 10.1101/2025.06.09.25329270

**Authors:** Xiaotong Yang, Wenting Liu, Zhixin Mao, Yuheng Du, Cameron Lassiter, Fadhl M. AlAkwaa, Paula A Benny, Lana X Garmire

## Abstract

Preeclampsia (PE) is a severe pregnancy complication that threatens maternal and neonatal health. Previous epigenome-wide association studies (EWAS) on PE have produced inconsistent results, possibly due to inadequate adjustment for confounders.

Here, we analyzed DNA methylation changes in cord blood from newborns affected by PE, using a multi-ethnic cohort from Hawaii. We comprehensively adjusted for clinical variables (maternal age, BMI, parity) and estimated cell proportions. Additionally, we re-analyzed two public datasets with similar adjustments and conducted a meta-analysis combining all three datasets to increase statistical power. To further address confounding by gestational age, we also included idiopathic preterm samples as controls.

After adjusting for cell type proportions and clinical characteristics, all previously reported significant CpG methylation changes associated with severe PE disappeared across our data, the two public datasets, and the meta-analysis. This result remained even after including idiopathic preterm samples. Instead, severe PE was associated with shifts in CD8T and natural killer (NK) cell proportions. We validated this lack of CpG changes using multiple published cord blood methylation datasets. Moreover, we observed that gestational progression itself is accompanied by significant changes in granulocyte, nRBC, CD8T, and B cell proportions.

In summary, our study demonstrates that many previously reported DNA methylation changes in severe PE are artifacts caused by confounding factors such as cell type heterogeneity and gestational age. Severe PE is associated with changes in cell proportions rather than direct methylation alterations. These findings emphasize the importance of rigorous confounder adjustment in EWAS.

## Introduction

Preeclampsia(PE) is characterized by new-onset hypertension with proteinuria or one/more adverse conditions after 20 weeks of gestation^1^. PE is one of the leading causes of maternal and prenatal morbidities and mortalities, affecting 2-8% of pregnancies globally and around 3.1% in the US^2,3^.

PE manifests as a diverse syndrome with multiple subtypes: based on blood pressure, clinical findings, and degree of proteinuria, PE can also be classified into severe PE or mild PE. Severe PE poses a greater risk to maternal and fetal health and may involve different pathways than mild PE of similar onset time^4^. Based on the onset time, PE can be divided into early-onset PE(EOPE), which occurs before 34 weeks of gestation, or late-onset PE(LOPE), which occurs after 34 weeks. The complexity of PE poses additional challenges in understanding its root causes.

Numerous studies have been conducted to investigate the molecular mechanisms of PE^5^, exploring genetic^6^, epigenetic^7,8^, transcriptomic^9^, lipidomic^10^, and telomere length^11,12^ changes in PE patients. However, most studies focus on the placenta and maternal tissue, while PE was reported to affect offspring, increasing their risk of hypertension, cardiovascular diseases and neurological development disorders^13^. The theory of the utero origin of diseases proposes that many chronic diseases are deeply rooted in the fetal stage, the very early phase of human development^14,15^. As the epigenome is both inheritable and prone to alteration by diseases, it is a plausible link mediating the effect of PE on offspring. Epigenome-wide association study or EWAS, identifies potential disease-associated DNA methylation markers and has been attempted by different studies to investigate if PE affects offspring ^16–20^. However, these studies did not reach coherent conclusions on the association between PE and cord blood DNA methylation profile.

Ching et al. first found significant global hypomethylation changes in cord blood affected by EOPE using the EWAS approach^16^. However, in the same year, Herzog et al. reported global hypermethylation in cord blood affected by EOPE compared to those in other preterm birth^17^, opposite to Ching et al. Later, Gao et al. claimed significant hypermethylation affecting the expression of AVPR1a, OXTR, and PKCB in preeclamptic umbilical veins^18^. Knihtila et al.^20^ found differentially methylated CpGs in preeclamptic cord blood associated with the cardiovascular pathway. Different from the above individual studies, Kazmi et al. conducted a large-scale meta-analysis (135 PE cases and 2084 controls) and reported 26 new significant CpG sites not previously associated with PE after adjusting for cell types and certain clinical factors. However, their results did not adjust for the gestational age, the major confounder of preeclampsia^19^. One possible reason for the inconsistency in previous studies may be due to the overlooked confounding effect: many of these earlier studies didn’t adjust for all potential confounders from cell-type heterogeneity^16,18,20^ and from important clinical variables such as gestational age(GA)^19^. A new study with good experimental design and rigorous statistical analysis is thus necessary to assess if these confounding variables truly contributed to the discrepancy, and detect the real association between PE and methylation patterns without such interference.

Cord blood consists of many diverse cell types, each with a distinct epigenome profile as defined^21,22^. Thus, the varying cell types in each sample can affect the overall DNA methylation profile at the bulk level^19^. It is therefore essential to account for such heterogeneity, to improve the accuracy and sensitivity, and avoid biased conclusions of EWAS biomarker detection. Particularly, to ensure that any differences in DNA methylation are due to confounding factors, the analysis needs to be adjusted for cell proportions. Moreover, if essential clinical data such as GAs are available (as they should be), then the analysis needs to be adjusted for the important clinical variables as well. In this study, we pay special attention to these issues to seek a plausible epigenomic association between severe PE and cord blood of offspring from PE patients.

## Materials and Methods

### The Hawaii Biorepository (HiBR) cohort

The umbilical cord whole blood DNA samples were obtained from HiBR, which was approved by IRB #CHS23976. HiBR collected placenta, maternal, and cord blood samples from deliveries at Kapiolani Women and Children’s Hospital from 2006 to 2013. It is one of the largest research tissue repositories in the Pacific region, containing specimens from more than 9250 mother-child pairs at the time of sample collection. The repository obtains informed consent from women post-partum to donate their placenta, umbilical cord, and excess cord and maternal blood (routinely collected for care purposes). Umbilical cord samples were collected immediately after delivery. Severe PE was characterized by OBGYNs at Kapiolani Medical Center as sustained pregnancy-induced hypertension(systolic/diastolic blood pressure >= 140/90) with urine protein and/or organ dysfunction.

This is part of a parental study to investigate the multi-omics biomarkers for severe preeclampsia and how the mothers and their female offspring are protected against breast cancers later in life, using the previously biobanked samples in the Hawaii Birth Repository (HiBR)^10,12^. The parental study includes women with severe PE who delivered singletons and were matched 1-to-1 by healthy PE-free deliveries based on maternal age, ethnicity, and pre-pregnancy BMI. For this nested cord blood study, we included those who delivered female babies and had cord blood samples remaining in the HiBR. We evaluated sample integrity, purity and concentration on the Nanodrop and removed samples of low quality. Variables such as maternal age, ethnicity, BMI, gestational age, parity, and smoking status were recorded as potential confounders. Patients with unknown ethnicity were excluded. This cord blood cohort contains 24 women with severe PE and 38 PE-free healthy controls. The demographic and clinical information of the patients was collected and analyzed to identify any potential confounding effects.

### Additional Cohorts

To validate the observations in the HiBR cohort, we did an exhaustive search among published work of cord blood (including PBMC) DNA methylation in association with PE^16,17,23^. We were able to obtain DNA methylome Illumina 450k data from the following studies: (1) Ching et al.^16^ with 12 PE cases and 8 controls, along with 6 clinical variables available, including maternal age, maternal BMI, maternal ethnicity, gestational age, baby gender, and baby birth weight. (2) Herzog EM et al.^17^ with 23 severe PE samples (including 10 early-onsets and 13 late-onsets) and 25 control samples, but no clinical variables available (GSE103253). and (3) cord blood leukocytes from Kashima K et al.^23^ with 20 PE cases and 90 controls, as well as 7 clinical variables available, including maternal age, maternal BMI, maternal smoking before pregnancy, parity, gestational age, baby gender, and delivery (GSE110828). We were unable to obtain data from the study by Kazmi et al. ^19^ as the data are not publicly available. We obtained the raw data from Ching et al. The data was filtered, normalized, and then corrected for batch effects by slide and array, using the R package “ChAMP”. For Herzog EM et al and Kashima K et al datasets, we directly used the beta matrix deposited to GEO.

To disentangle the effect of PE and small GA, we also included cord blood samples from Fernando et al.(GSE66459)^24^. This dataset includes 11 idiopathic preterm birth samples and 11 full-term samples. All samples from this cohort are PE-free. We directly used the normalized matrix deposited to GEO.

### Sample preparation

Umbilical cord blood samples were collected immediately after delivery. To prepare for DNA extraction, we first added three volumes of RBC Lysis Solution to one volume of clotted blood, which was then vortexed and incubated on a shaker for 15 minutes at room temperature. The sample was then centrifuged to pellet white blood cells and clot particulates, and the supernatant was carefully poured into a waste bucket. The pellet was resuspended in an additional volume of RBC Lysis Solution and incubated again for 15 minutes. After another centrifugation step, the supernatant was carefully removed, leaving behind 200 µL of residual liquid. The pellet was then vigorously resuspended in the residual liquid before being combined with a master mix of Cell Lysis and Proteinase K Solution. The mixture was vortexed and incubated at 55°C until homogenous, with intermittent vortexing to facilitate digestion. Once homogenous, the samples were subjected to DNA purification on the Autopure Machine following the manufacturer’s instructions.

### DNA extraction and methylation profiling

DNA was extracted from prepared cord blood samples by HiBR using AllPrep DNA/RNA/Protein Mini Kit (Qiagen, USA) according to the manufacturer’s instructions. We obtained pre-extracted genomic DNA of whole cord blood samples from the HiBR and conducted DNA Illumina EPIC Beadchip assays through the University of Hawaii Cancer Center Genomics Core. Case and control samples are interleaved on the plate to ensure the sample group is independent of batch effects.

### DNA methylation data pre-processing and quality control

We used the R package “ChAMP” for data pre-processing (**Supple Fig. 1).** We first filtered probes using the following criteria sequentially: (1) removing probes with a detection p-value above 0.01 (7,941 probes); (2) removing probes with a bead count <3 in at least 5% of samples (27,731 probes); (3) removing probes with no matched CpG sites (2,673 probes); (4) removing probes that align to multiple locations (8,248 probes). During the quality control step, we removed 1 control sample with a distinct beta density distribution (**Supple Fig. 2A, 2B**). We normalized the remaining samples using BMIQ methods^25^, and corrected for batch effects ( slide and array) using the ComBat algorithm embedded in “ChAMP”. We used the singular vector decomposition (SVD) heatmap to verify the effectiveness of batch removal (**Supple Fig. 2C, 2D**). SVD deconvolution reconstructs the original methylation matrix into principal components(PC), ordered from the most informative to the least. Colors in the SVD heatmap (**Supple Fig. 2C, 2D**) represent the correlation of each variable to each PC with a deeper color indicating a stronger correlation. After batch correction, the correlations between the top PC and batch variables (slide and array) were removed, suggesting a successful correction (**Supple Fig. 2C, 2D**). The preprocessed data matrix contains 62 samples and 819,325 probes. We converted the original methylation intensity (beta) to M-values using “beta2m” function from “lumi” package to reduce heteroskedasticity^26^, where M-values are defined as the log2 ratio of the beta value of each probe.

### Cell-type deconvolution in umbilical cord whole blood (CB)

Bulk-level DNA in umbilical cord whole blood (CB) includes at least 7 most common blood cell types: granulocytes, B cells, CD4T, CD8T, monocytes, natural killer cells (NK), and nucleated red blood cells (nRBC). Each sample may have different compositions of the cell types above, thus needing deconvolution. We adopted the Houseman’s constrained projection(CP) algorithm^27^ and a combined cord blood cell type reference as recommended by Gervin et al ^22^. This combined reference includes 263 cord blood cell type signatures from 4 previous large studies^28–31^ and is used in many previous cord blood cell type estimation studies^32,33^. We used the resulting cell type proportions to adjust for confounding effects in the differential analysis step.

### Clinical confounders and source of variance analysis

We retrieved a total of 6 commonly reported clinical variables in cord blood EWAS study from the biobank, including maternal age, ethnicity, parity, BMI, delivery GA, and smoking status^19^. We imputed 3 samples (including 1 severe preeclampsia and 2 controls) with missing BMI using mean values of each sample group. We performed the source of variance (SOV) analysis on these clinical variables and previously estimated sample cell proportions to identify important confounding variables that need to be adjusted, as done before^10,34,35^. Specifically, we regressed each CpG site on all clinical confounders and applied two-way ANOVA to each regression. Next, we averaged the F-statistics for each variable across CpG sites and retained those with F-statistics greater than 1 (the value of the noise) for downstream differential analysis. This approach eliminates non-informative clinical variables and selects a linear model that provides the best overall fit.

### CpG-level epigenome-wide association analysis (EWAS)

We calculated the differentially methylated probes (DMP) between severe PE cases and controls by fitting linear regression with empirical Bayes moderated statistics on each probe. The p-value of PE was adjusted with Benjamini-Hochberg (BH) adjustment (threshold of 0.05). We included study participants’ GA (GA), BMI, parity status, ethnicity, and methylation-derived cell compositions in the linear model to remove the confounding effects, and compared the result with that without confounding adjustment using “limma” package^36^. We defined hypermethylated CpGs as significant CpGs with positive log2-transformed fold change (logFC) and hypomethylated CpGs as significant CpGs with negative logFC, respectively. We used volcano plots to illustrate the global DNA methylation changes between the cases and controls.

### Meta-analysis using three datasets

We also combined the three cohorts(in-house, Ching et al., Herzog et al.) and conducted a meta-analysis to improve the test power and produce more robust results. We did not include data from Kashimi et al. because there was no significant cpg before the confounder adjustment.

To harmonize the datasets, we first filtered the raw in-house EPIC data and raw 450k data from Ching et al. individually, then merged them based on overlapping CpG sites and normalized the dataset. Next, we combined it with the normalized beta matrix from Herzog et al., downloaded directly from GEO. To harmonize the datasets, we applied the combat function to remove batch effects while preserving sample group information. We calculated the differentially methylated probes between severe PE cases and controls using limma and plotted the volcano plot. We then estimated the cell proportion of the merged data using Houseman’s CP method and the cell-type reference by Gervin et al. Lastly, we calculated differentially methylated probes again, with adjustment to the sample group and estimated cell types and plotted another volcano plot.

### Including idiopathic preterm samples to decouple the effect of PE and small gestational age

Many PE patients are delivered preterm to avoid severe maternal complications, resulting in an inevitable correlation of PE cases and smaller gestational age. To decouple the effect of PE and GA, we include another study, Fernando et al. (GSE66459)^24^. The dataset contains DNA methylation data of 11 idiopathic preterm and 11 term samples processed with the Illumina 450K Human methylation bead chip array. We merged our in-house data with Fernando et al. data by common cpgs, and harmonized the two datasets with the Combat algorithm, same as in the meta-analysis above. We then computed the differentially methylated probes with adjustment of gestational age, baby sex and estimated cell proportion again using the limma package. We examined and corrected for potential bias and inflation using the empirical null distribution from the “bacon” R package^37^.

### Differential methylated regions(DMR)

To identify the differentially methylated regions(DMR), we used the bumphunter method from R package “bumphunter”. The result is adjusted for the same clinical variables and cell type proportions as the CpG-level differential analysis. We used the FWER method to adjust the P-values of each DMR and used adjusted p-value <0.05 as the cutoff for significant DMRs.

### Gene-level EWAS

We further examined the methylation signal difference between severe PE and controls at gene and pathway levels. We annotated the CpGs^38^, selected those located on the promoter region and aggregated the methylation signals of all CpGs within a gene promoter by taking the geometric mean. Then we compared the aggregated methylation signals between severe PE cases and controls, using linear regression with empirical Bayes moderated statistics. Additionally, we looked for pathways associated with promoter region methylation differences using the R package pathifier^39^. The *Pathifier* algorithm calculates a pathway deregulation score (PDS) for each sample and each pathway. We compared the pathway PDS scores in case and control samples adjusted for the same confounders as the probe-level analysis; the result p-values were adjusted with Benjamini-Hochberg FDR (threshold p-values 0.05).

### Software Usage and Code Availability

All analysis was done using R 4.1.2^40^. Specifically, we used “ChAMP” (version 2.24.0) for data preparation^41^, “limma” for differential methylation analysis^36^, “EpiDISH” (version 2.10.0) for cell-type deconvolution^42^, and “IlluminaHumanMethylationEPICanno.ilm10b4.hg19” for data annotation^38^. All codes are available at https://github.com/lanagarmire/CB_DNAm_PE.

## Results

### Overview of Study Design and Cohort Characteristics

The overview of the study design is illustrated in **Fig. 1**. We obtained whole cord blood samples from 24 severe PE cases and 39 controls collected at the Hawaii Biorepository (HiBR) from January 2006 and June 2013. The maternal demographic and clinical characteristics are shown in **Table 1**. There is no significant difference (P > 0.05) in maternal age, parity, BMI, ethnicity, and smoking status. Noticeably, the severe PE cases have earlier delivery GA compared to healthy controls (P = 3.66E-06), as the clinical management of severe PE often demands early delivery to avoid severe maternal morbidities. We obtained pre-extracted genomic DNA of whole cord blood samples from the HiBR and conducted DNA Illumina EPIC Beadchip assays through the University of Hawaii Cancer Center Genomics Core. We conducted bioinformatics pre-processing of the DNA methylation following standard steps in R package “ChAMP” (**Supple Fig. 1**). Briefly, we first filtered out probes of bad quality, then examined the methylation level distribution of each sample and removed 1 control sample with abnormal distribution (**Supple Fig. 2A, 2B**). Lastly, we normalized the data and removed the batch effect between arrays (**Supple Fig. 2C, 2D**). The remaining preprocessed data matrix contains 62 samples and 819,325 probes (see **Methods**).

**Figure 1:**
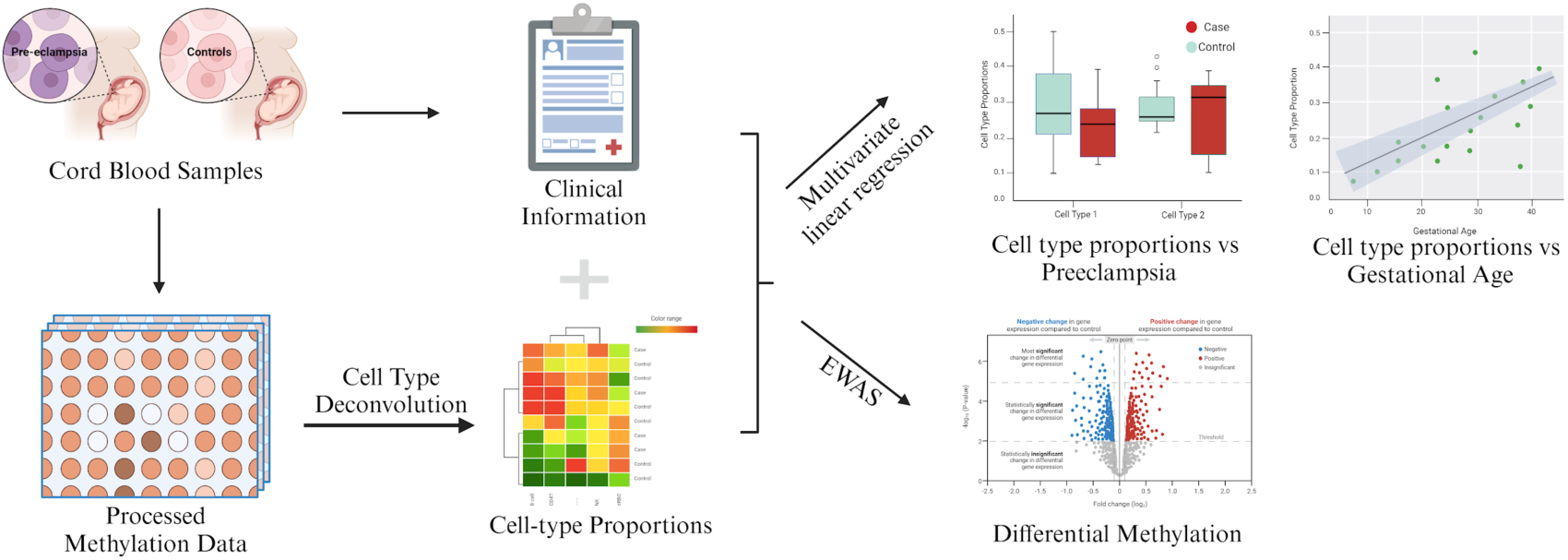
Study Overview and Experiment Design. The entire data analysis procedure is outlined in this workflow, which incorporates methods that account for clinical confounding and cell-type confounding. Created with BioRender.com

**Table 1:** Patient Characteristics.

| Variables | PE Cases<br>(n = 24)<br>mean (sd) | Controls<br>(n = 38)<br>mean (sd) | P-value |
| --- | --- | --- | --- |
| Maternal Age (Years) | 28.75 (5.88) | 27.24 (6.35) | 0.34 |
| Parity | 1.54 (1.41) | 1.57 (1.78) | 0.93 |
| BMI | 32.24 (9.38) | 27.75 (9.20) | 0.07 |
| Smoker (n) | 6 | 8 | 0.96 |
| Gestational Age (Weeks) | 35.58 (2.90) | 39.16 (0.92) | 3.66E-06 |
| Ethnicity (n) |  |  | 0.60 |
| Asian | 12 | 21 | - |
| Caucasian | 3 | 7 | - |
| Pacific Islander | 9 | 10 | - |

\* Numeric variables are compared with t-test; \* Categorical variables are compared with the Chi-square test.

### Associations between cord blood cell types and severe PE

To learn the association between severe PE and cord blood cell composition, we performed cell-type deconvolution using the cord blood cell type reference as recommended by Gervin et al^22^ and Houseman’s constrained projection (CP) deconvolution algorithm^27^. The estimated cell-type proportions show large variations across samples, especially for granulocyte, nRBC, and CD4T cells (**Fig. 2A**). Granulocyte proportions appear significantly lower (t = −5.60, P <2.80e-06) in the severe PE group compared to the control group, while B cell (t = 2.09, P < 0.05), nRBC cell (ꞵ= 4.17, P<0.001) and CD8T cell (ꞵ=3.45, P<0.01) proportions seem significantly higher in cases (**Fig. 2B**).

**Figure 2:**
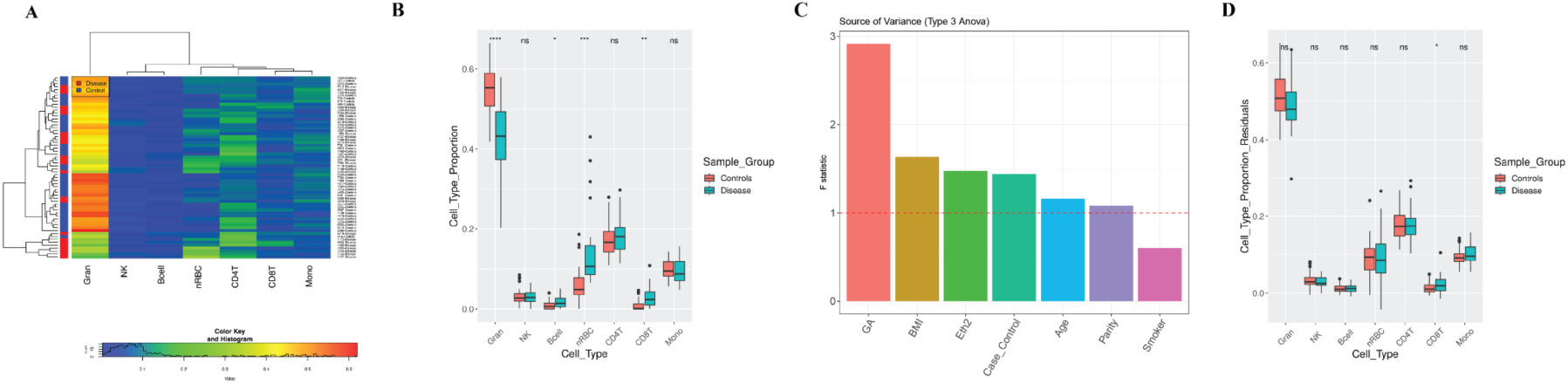
Cell types in samples. (A) Heatmap displaying estimated cell-type proportions among 62 samples (including 24 PE cases and 38 controls), the colors indicate the relative proportions of cell types, with red indicating a higher proportion and blue indicating a lower proportion. (B) Side-by-side boxplots displaying cell-type proportions in PE cases vs. controls before adjusting for clinical variables. An asterisk (*) is used to indicate a significant difference by using Multiple Linear Regression (MLR) between the case and control groups (p-value < 0.05), while “ns” is used to indicate a non-significant difference. (C) The Source of Variance (SOV) analysis of clinical covariates. Confounding factors were identified by considering variables with an F-mean value greater than 1. (D) Side-by-side boxplots displaying cell-type proportions in PE cases vs. controls with p-values from multiple linear regression of cell proportion on PE and all confounders identified in (C).

However, the apparent differences in cell proportions in cases vs controls could be very well due to other reasons (eg. GA) rather than the severe PE condition itself. To confirm this speculation, we conducted the source of variance (SOV) analysis of cell type compositions on the clinical variables and ranked them by F-statistics. A variable with an F-statistic bigger than 1 (the error term) is considered a significant contributor to cell proportion variations. As shown in **Fig. 2C**, in addition to severe PE, GA, maternal BMI, and ethnicity also contribute significantly to cell type heterogeneity. GA (F-statistic = 9.30) and maternal BMI (F-statistic = 3.10) rank higher than severe PE (F-statistic = 2.41), with GA being the predominant influencing factor. With such caution, we calculated the association between cell proportion and severe PE again with linear regression. This time, we include GA, maternal age, ethnicity, BMI, and smoking status as covariate factors. For comparison, we plot the cell proportions in severe PE vs. controls and reported p-values of PE from linear regression with other clinical covariates (**Fig. 2D**). The previously observed differences in B cell and nRBC proportions now disappear. CD8 T-cell proportions, however, continue to be significantly higher in severe PE cases (ꞵ=0.018, P=0.017). Granulocyte proportions show a trend of decrease in severe PE cases compared to controls, although not statistically significant (ꞵ=-0.047, P=0.059). The detailed linear regression results of cell proportion on clinical data can be found in **Supplementary Table 1**. The previous differences observed in B cells and nRBCs are indeed due to different gestational ages that confounded PE cases. In all, these results show that certain cord blood cell proportions vary among newborns and it is important to adjust for potential confounding before interrogating the association with severe PE.

### Lack of association between cord blood DNA methylation and severe PE

Considering that most previous cord blood EWAS studies overlooked the adjustment of cell types or other clinical covariates (such as GA) within their samples, we further investigated the impact of these factors on the differential methylation analysis. We conducted the source of variance (SOV) analysis of the DNA methylation matrix on cell type proportion and clinical variables. Strikingly, all cell-type composition variables show the strongest and most dominant explanatory power of variation in the methylation data (**Fig. 3A**), ranking even higher than the severe PE condition itself. After the cell proportions, severe PE case/control, GA, maternal age, parity, and ethnicity also have larger F-statistics than the error term (F-statistics=1), in descending order. Therefore, in the downstream analysis of differentially methylated CpGs, we adjusted for these confounding variables.

**Figure 3:**
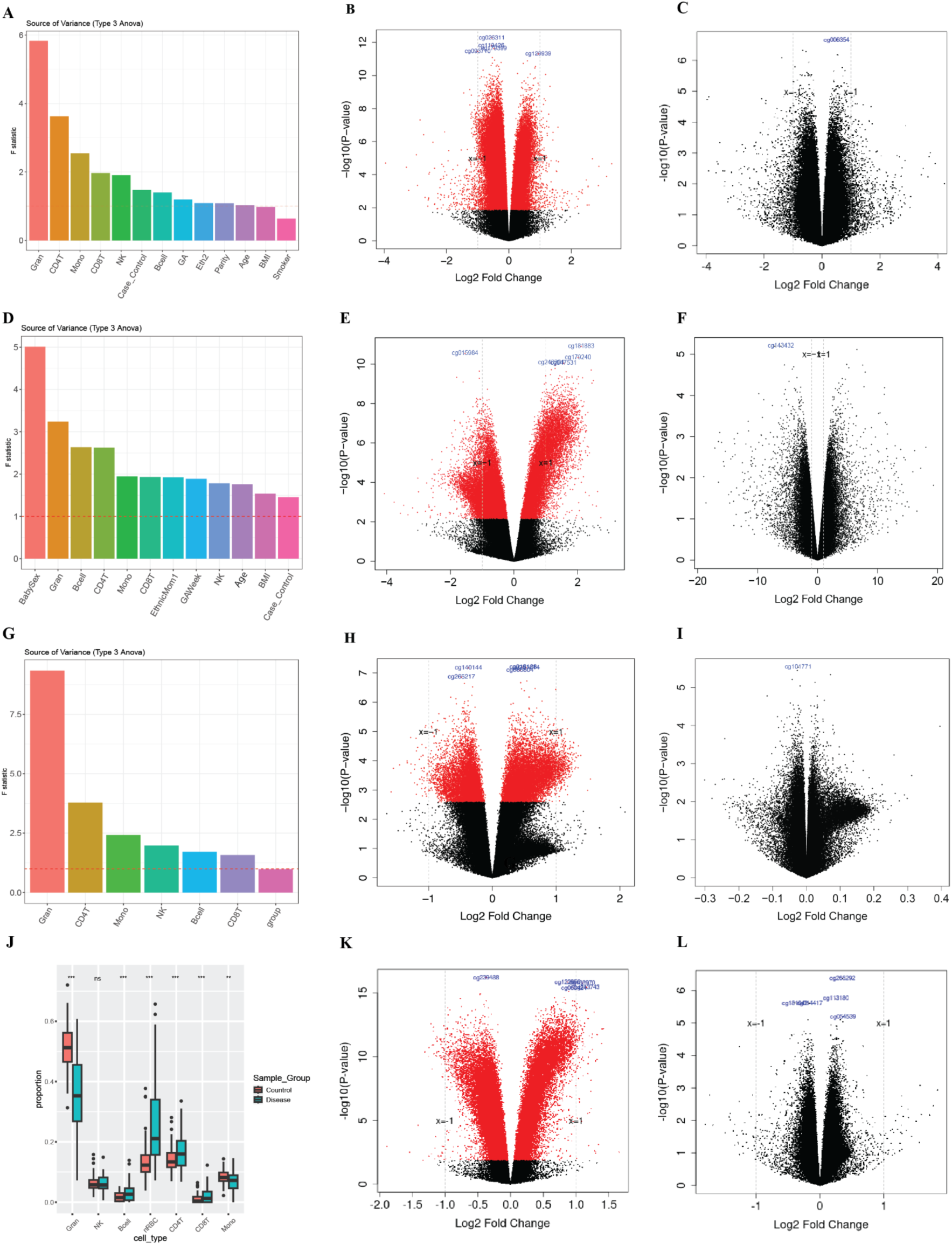
PE is not associated with significant changes in cord blood DNA methylation, after confounding adjustment, using data from HiBR cohort (A-C), Ching T et. al (D-F), Herzog EM et al (G-I) and meta-analysis combining the three cohorts (J-L). (A, D, G) The Source of Variance (SOV) analyses were conducted on cell types and clinical variables whenever available. Confounding factors were identified by considering variables with an F-mean value greater than 1. (B, E, H, K) The volcano plot of the differential methylation analysis results without confounding adjustment. The x-axis represents log fold change between severe PE and controls; the y-axis is negative log-transformed p-values after BH adjustment. The red dots are differentially methylated probes (DMP) associated with severe PE after BH adjustment, whereas the black dots represent non-significant probes. (C, F, I, L) The volcano plot after adjusting for all confounding factors. (J) The cell proportion distribution in the combined 3 datasets, separated by sample groups.

As a comparison, we first conducted differential methylation analysis on severe PE without adjusting for any clinical confounders or cell-type proportions. The analysis reveals a global hypomethylation pattern (**Fig. 3B**). We identified 229,730 differentially methylated CpGs with adjusted p-values less than 0.05. Among these CpGs, 184,102 exhibited hypomethylation, while 45,628 displayed hypermethylation. However, when we redid the differential methylation analysis after adjusting for cell type heterogeneity and patient characteristics, all the CpGs differentially methylated above were no longer significant (**Fig. 3C**). Since previous studies reported that maternal smoking significantly affects DNA methylation^43^. To consider this we included the variable “smoking” despite its smaller effect on data variance. This did not change the observation of a lack of significant CpGs from the cord blood associated with severe PE (**Supple. Fig 3)**. Additionally, we conducted differentially methylated region (DMR) analysis, which also suggests no significant association with severe PE when adjusted for the same confounders (**Supplementary Table 2**). We extended the differential methylation analysis to the gene level, by aggregating the CpGs located on gene promoters as the representation of promoter-level methylation (see **Methods**). Before adjusting for confounders, we detected 4,767 differentially methylated genes. However, upon adjusting for both clinical variables and cell types, none of the genes exhibited statistical significance. Similarly, we conducted a differential methylation analysis at the pathway level, employing the Pathifier algorithm (see **Methods)**. Before the confounding adjustment, we detected 200 significant pathways; after accounting for confounders, none of the pathways remained significant. In conclusion, we found that the observed DNA methylation variation among the whole cord blood samples is primarily associated with cell type differences rather than severe PE.

To confirm this surprising finding that contradicts all previous EWAS studies on cord blood samples associated with PE, we re-analyzed all other available public whole cord blood (or PBMC) DNA methylation datasets associated with PE samples, from Ching et al.^16^, Herzog EM et al.^17^, and Kashima K et al.^23^. We estimated the cell type proportions the same way, using Houseman’s CP algorithm and the new combined cord blood reference recommended by Gervin et al^22^. We conducted the SOV analysis by considering cell proportions in both studies and clinical variables whenever available (for Ching et al). SOV shows cell type proportions, GA, maternal age and PE as significant confounders, as they have F-statistics > 1 (**Fig. 3D, 3G**). For the dataset of Ching et al, we used the original analysis pipeline that did not consider clinical confounding and reproduced the differential methylation results earlier, which reported 68,458 significant CpGs (**Fig. 3E**). However, once we adjust for the cell types and other clinical confounders, there are no longer significant CpGs (**Fig. 3F).** For the newborn umbilical cord blood dataset of Herzog EM et al., we conducted a differential methylation analysis as well. Without adjusting for any clinical confounders or cell-type proportions, we obtained 24,597 significant CpGs (**Fig. 3H**). Again, once we adjusted the cell type proportions (all seven major whole cord blood cell types: monocytes, CD4T, natural killer, granulocytes, nRBC, B cell, CD8T) there is no significant CpG remaining (**Fig. 3I**). For Kashima K et al. data, we found no significant CpGs even before cell type adjustment (**Supple. Fig. 4**). Thus, using all other three available cord blood datasets, we confirm there indeed is a lack of association between cord blood DNA methylation and severe PE.

To increase the statistical power, we further conducted a meta-analysis by combining our in-house data, data from Ching et al. and Herzog et al.. The three datasets were processed and harmonized as described in the Method section. Again, the differential methylation result of combined data shows many significant CpGs are associated with PE before confounder adjustment (**Fig. 3I**), but are insignificant after adjusting for cell proportions (**Fig. 3L**). Additionally, to best decouple the effect of PE and early gestational age, we included the idiopathic preterm samples from Fernando et al.^24^. The dataset has 11 preterm and 11 full-term samples whose cord blood DNA was processed with Illumina 450k beadchips. We merged them with our HiBR data and computed the differentially methylated CpGs (see **Methods**). Consistent with previous conclusions, we found no significant difference between PE cases and non-PE controls, after adjusting for cell proportion, baby sex and gestational age (**Supple. Fig. 5**). We thus conclude that there is a lack of association between cord blood DNA methylation and severe PE.

### Association between cord blood cell type and GA

Our earlier analysis shows that estimated cell proportions in cord blood are mostly correlated with GA (**Fig. 2C**). We thus conducted a more in-depth analysis. The most noticeable correlation comes from granulocytes, whose proportions increase from around 25% in week 32 to over 50% in week 40, with p<2.11e-11 (**Fig. 4A**). The proportions of monocytes also significantly increase as gestation progresses, after adjusting for other variables (p=0.019). On the contrary, B cell, CD8T, and nRBC significantly decrease along the gestation (p=4.99e-4; p=2.08e-3; p =9.13e-10). We also plotted the trends of cell type proportions related to GA per sample group, by merging our HiBR and Fernando et al.’s data (**Fig. 4 B**). These trends of cell proportions are mostly the same in the case and the control group, except for monocytes, which show a potential interaction effect between PE and gestational age.

**Figure 4:**
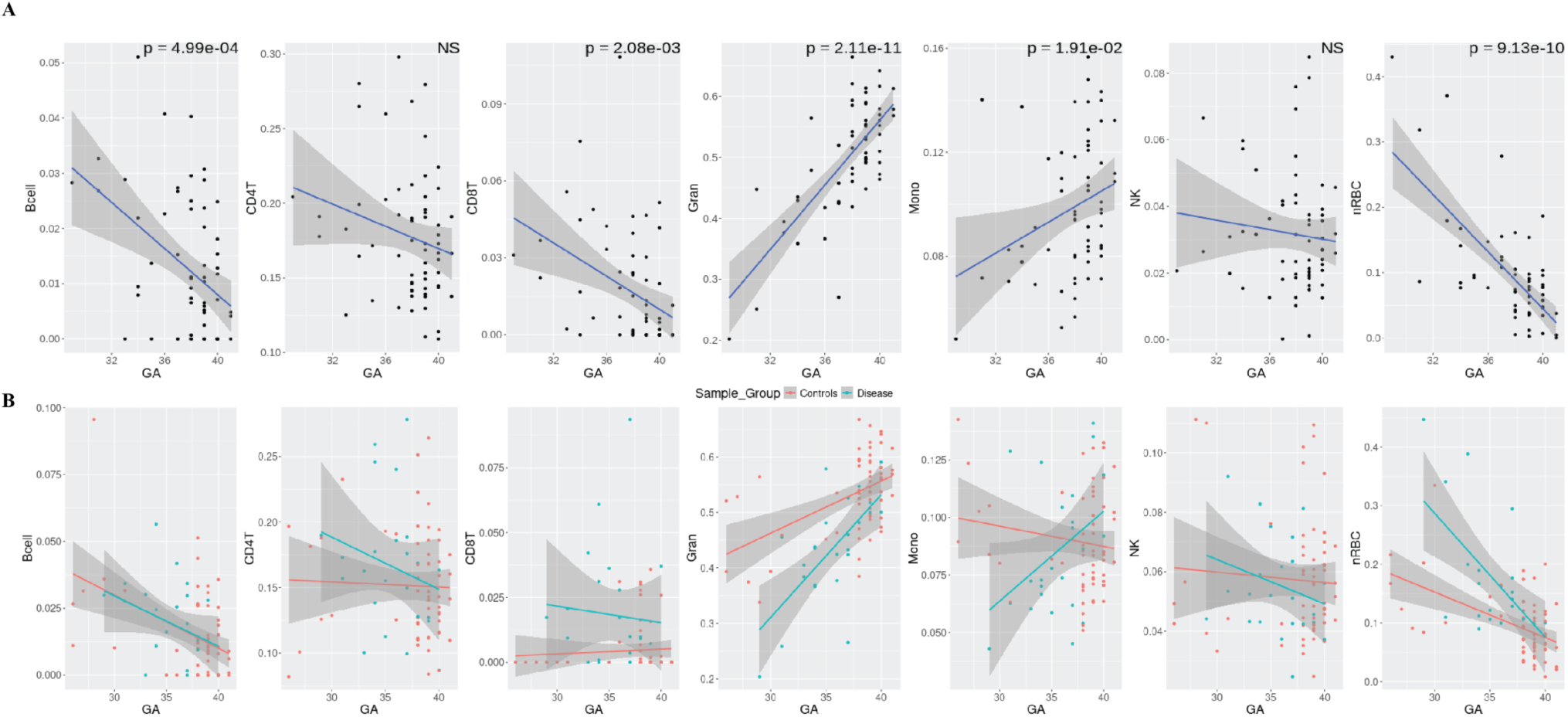
Cell type proportions in relationship to the gestational age. Scatter plots (A) depict the proportions of each cell type in cord blood from all samples, along with gestational age. The reported p-value measures the relationship between GA and each cell type, with a threshold of p-value < 0.05. (B) Scatter plot of estimated cell proportions from merged HiBR data and those from Fernando et al., by gestational age.

Furthermore, we validated the trends of cell type proportion through GA using another public cord blood peripheral blood mononuclear cell (PBMC) Illumina HumanMethylation450 BeadChip methylation dataset (GSE110828), which comprises 20 PE cases and 90 non-PE controls^23^. Both the case and control groups include large percentages of preterm samples, with the delivery GAs ranging from 26.14 to 38.14 weeks in cases and 23.00 to 41.29 weeks in controls. We deconvoluted the PBMC cell types using the same combined cord blood reference. To ensure comparability of cell proportions between PBMC and whole blood, we eliminated granulocytes, which are unique to whole blood and recalibrated the weights of the remaining cell types to sum to one. In both PE (**Fig. 5A**) and control samples (**Fig. 5B**), we observed the same increasing trend for monocytes, and the same decreasing trend for CD8T, nRBC cells, and B cells. For NK cells, while both cohorts show consistent trends of decrease in the control samples with GA, the trend in PE samples is not conclusive, possibly related to the small sample size (n=20) in the other PBMC cohort. Further, to test if the cell proportion trends in relationship with GA are consistent between the two datasets, we performed linear regression of each cell proportions (y-variables), over GA, PBMC vs. whole blood dataset stratification, and the interaction terms between datasets and GA, with adjustment of other confounders for PE cases and controls separately. None of the interaction terms between datasets and GA turn out to be significant, as the p-values in **Fig. 5**. A non-significant p-value of the interaction term indicates the cell proportion trends are not statistically different between the case and control groups. This shows that the GA’s effect on cell proportions is consistent and independent of datasets.

**Figure 5:**
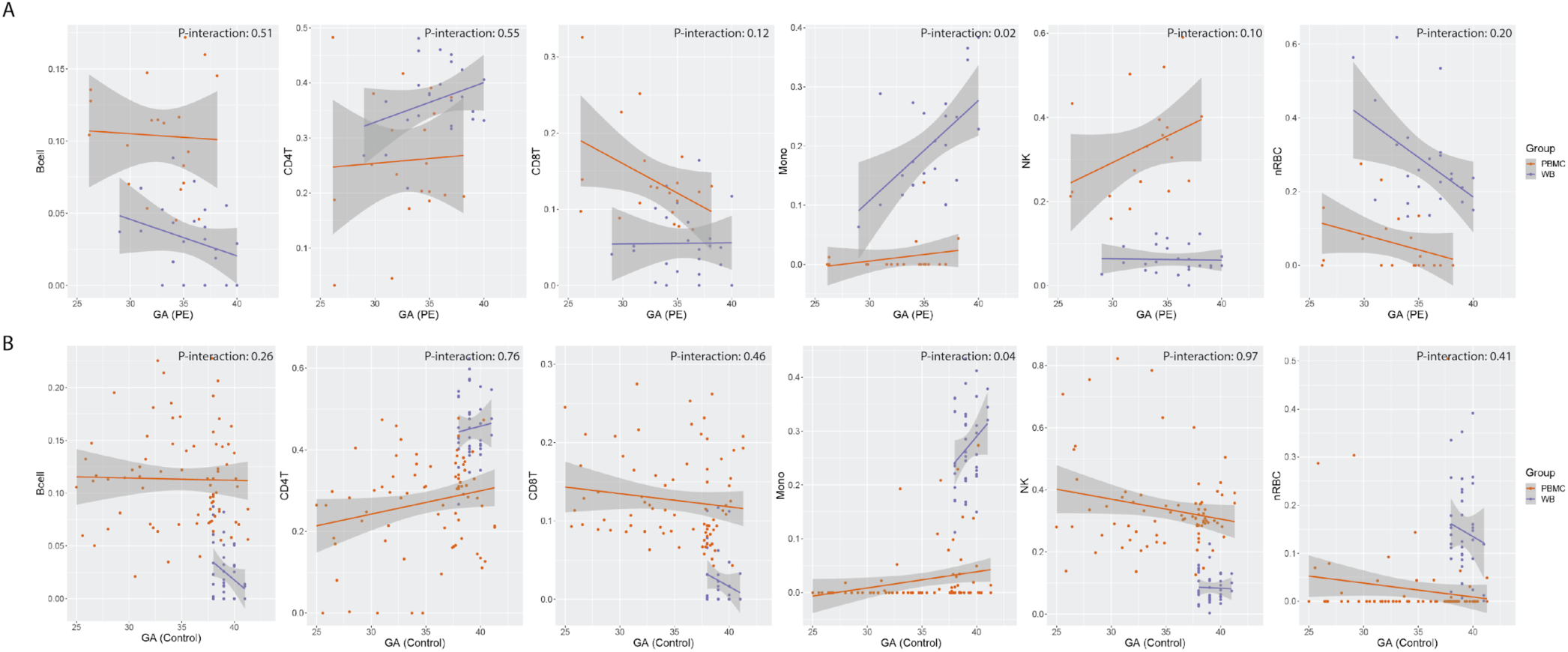
Cell proportions in relationship with gestational age are coherent in two different datasets. The scatter plots compare the cell-type proportions with gestational age in our data and Kashima et al.. Plots (A) display the comparisons within PE case samples for both studies, whereas plots (B) display the comparisons within control samples for both studies. The purple line in each plot represents the fitted cell proportions in our whole cord blood samples, while the orange line represents the fitted cell proportions in PBMC cord blood samples from Kashima K et al., another study. We test whether the cell proportion changes in the two datasets, using PBMC vs. the whole blood, are significantly different by linearly regressing cell proportions over GA, datasets, and the interaction between the datasets and GA. We report the p-values of such interaction terms. A non-significant p-value indicates the cell proportion trends are not statistically different in the two datasets.

## Discussion

In this study, we showed there is a lack of association between severe PE and DNA methylation level changes, in the cord blood samples of the offspring of these PE patients from multiple cohorts. We carried out the study with rigorous confounder adjustment for both cell type heterogeneity and clinical variables, especially gestational age. Interestingly, aligning with our observation, recent work by Campbell KA et al also showed the huge confounding effect of cell types on placenta gene expression changes associated with preeclampsia^44^. Despite the absence of CpG changes associated with severe PE, we observed noticeable variations in the CD8 T-cell proportions between severe PE cases and controls. We conclude that the primary impact of maternal PE on the offspring’s cord blood is not methylation alteration. Instead, severe PE manifests itself impact on the offspring of the affected mother by altering the proportions of some immune cell types in the blood. In particular, CD8T cells are significantly higher in the baby’s cord blood from the cases, after adjusting for other confounders such as GA, possibly due to the activation of the innate immune system of the babies from PE patients. Previous studies have well confirmed the activation of T cells in preeclamptic patients^45,46^. This observation is now expanded to their offspring, demonstrating the impact of PE. The mechanism for increasing CD8T cells in cord blood is of interest for future work.

Furthermore, we noticed generally consistent associations between cell type compositions in cord blood and GA in two independent cohorts, regardless of the existence of severe PE conditions. Granulocyte proportion showed the strongest quantitative changes along GA, agreeing with the previous findings that granulocyte in the fetus increases drastically in the last trimester of pregnancy^47,48^. On the contrary, the estimated proportion of nRBC in our study decreases drastically as GA increases, also consistent with previous findings^49–51^. Previously elevated nRBC was also found in preterm infants and infants with lower birth weight^52^, providing additional supporting evidence to our finding. Braid et al. also reported decreases in B cells in cord blood using methylation-derived cell proportion. One explanation is that the large increase of granulocytes in late gestation makes the proportions of other cell types smaller, not necessarily the absolute value. Another possibility is that preterm infants are experiencing higher inflammation levels, which leads to higher B cell proportion^47^. Taken together, these findings probe into the dynamic nature of cell type composition in cord blood during gestation, confirming the importance of cell type adjustment in methylation analysis.

Among all confounders in EWAS analysis, cell type heterogeneity is one of the most common and important confounders faced by researchers. Over the years, various cell-type estimation methods for bulk-level epigenetic data have been developed ^27,53^, making the assessment of cell-type confounding effects possible. In 2013, Liu et al. first reported a large reduction in differentially methylated probes related to Rheumatoid Arthritis after adjusting for cell type composition in whole blood^54^. Some later studies confirmed the effect of cell type heterogeneity on EWAS research in other tissues, such as breast tissue, saliva, and placenta tissue^32,44,55–57^, emphasizing the importance of adjusting for cell type heterogeneity in the EWAS of PE studied here. Kazmi et al. conducted a large meta-analysis on the association of PE and newborn DNA methylation and detected a small number of 26 significantly associated CpGs. However, they did not adjust for GA due to the concern of its confounding effect on PE^19^. We showed that GA affects the methylation pattern both directly and through cell proportion change, as reported before^58^; therefore, it should be adjusted. More importantly, the cell proportions in severe PE case groups are also affected by the disease itself, making them critical confounders in EWAS studies. Although several previous studies aimed to identify PE-related epigenetic biomarkers using cord blood samples^16–19^, the importance of adjusting for cell type heterogeneity was mostly (3 out of 4) overlooked. Using as many as four different cohorts, our investigation here shows that ignoring the cell type heterogeneity and gestational age may have contributed to biased EWAS associations with PE, as done previously by multiple studies.

Some caveats are worth mentioning for this study. First, all the cell types in the blood are computationally inferred, rather than experimentally measured. In theory, technology such as flow cytometry is likely more powerful for direct applications in EWAS or PE. However, for retrospective studies such as this one, where only whole blood DNA was available, or for historically archived bulk DNA methylation data, computational deconvolution is the only viable option. Also, infections such as chorioamnionitis, which unfortunately is not part of the collected clinical variables here, may also influence the association between GA and cell proportions by triggering a surge of neutrophils. However, given the small proportions of chorioamnionitis, this might not be a major issue. In addition to the confounders we already included, genetic changes such as polymorphism, genetic relatedness, and structural variation may also play roles^59^. Our attempt to remove SNP-overlapped CpGs should mitigate these issues to some degree. Future EWAS studies should carefully consider these confounding factors to ensure the scientific rigor of research. This study focuses on cord blood DNA methylation changes, but the health of the fetus often depends on the degree of placental damage. We did not detect significant differences in the inferred proportion of endothelial cells between the control and PE groups using the DNA methylation data from paired placental samples. Future follow-up histopathological investigation into the maternal decidua may examine vascular malperfusion, but beyond the scope of this study. Lastly, the cord blood samples studied here are modest in size (n = 62). The subsequent meta-analysis in Figure 3L has a total of 129 (PE = 58) cord blood samples. We could not exploit the data in Kazmi et al. due to the lack of open access^19^. It will be highly interesting to combine these data together for re-analysis with statistical rigor in the future.

## Conclusion

In summary, we could not find the evidence for significant CpG methylation changes in EWAS analysis in association with severe PE, after adjusting for cell type heterogeneity and clinical variables such as GA. Instead, severe PE is associated with significant changes in several cell proportions in the cord blood. Additionally, many cell type proportions change drastically as pregnancy progresses.

## List of abbreviations

PE: preeclampsia
EWAS: epigenome-wide association study
GA: gestational age
PBMC: peripheral blood mononuclear cell
NK: natural killer
nRBC: nucleated red blood cell

## Data Availability Statement

All supporting DNA methylation data has been deposited in Gene Expression Omnibus (GEO). The associated accession number will be added upon approval.

## Author Contributions

LXG conceived this project and supervised the study. XY and WL contributed equally to data analysis, result generation, and manuscript writing. ZM and YD assisted in the data processing. CL, FMA, and PAB contributed to sample collection, coordination and the experimental design of the DNA methylation. All authors have read, revised and approved the manuscript.

## Acknowledgment

We thank the Genomics Shared Resources of the University of Hawaii Cancer Center for performing the methylation assays.

## Materials & Correspondence

Correspondence to Lana X Garmire

