## Supplementary Tables and Figures for "Cell type proportions rather than DNA methylation in the cord blood show significant associations with severe preeclampsia"

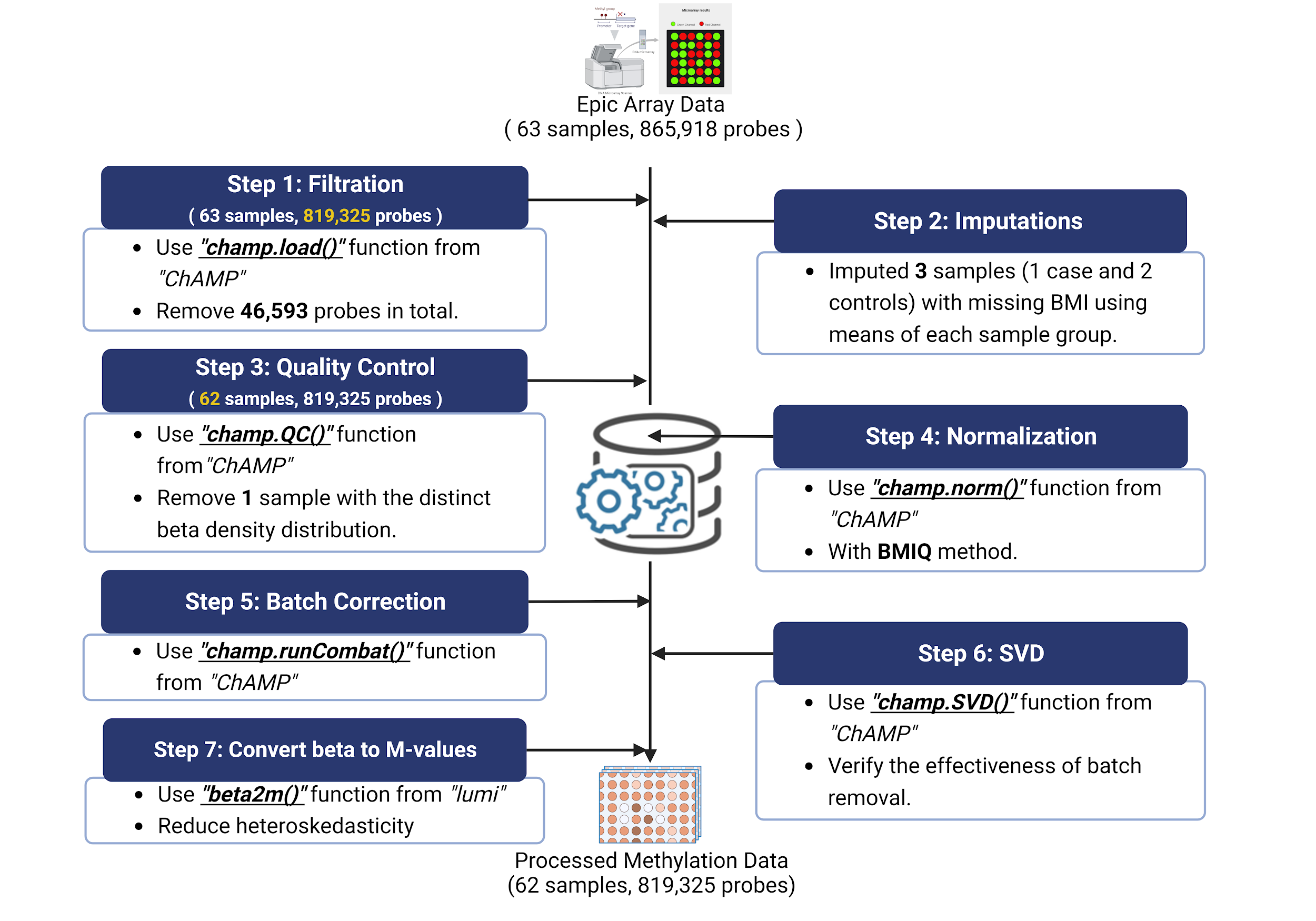

**Supplementary Figure 1:** Data Processing Workflow. The complete data pre-processing procedures consisted of filtration, imputation of missing values, quality control checks, normalization, batch correction, singular value decomposition analysis, and conversion of beta values to M-values.

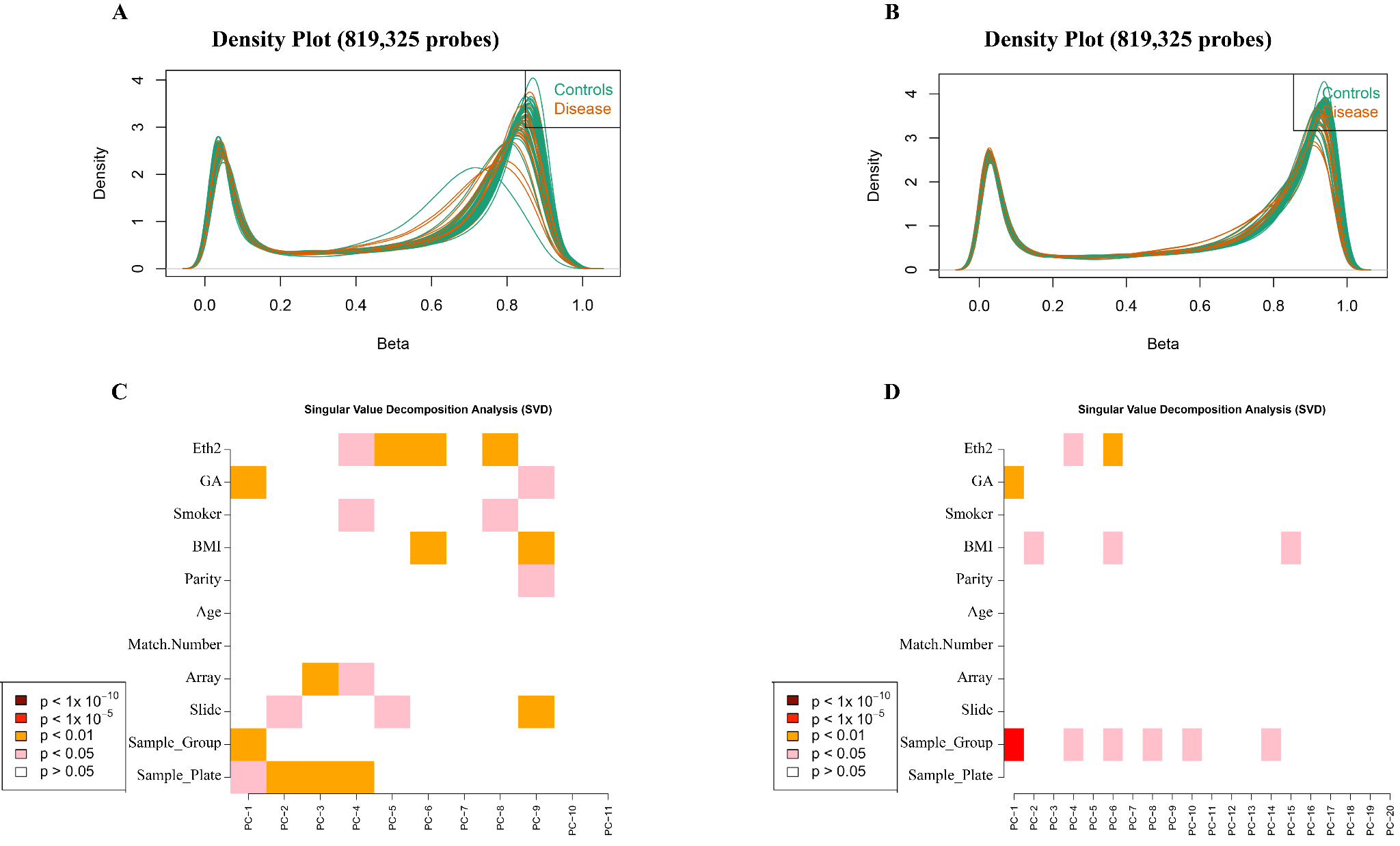

**Supplementary Figure 2: Data Quality Control.** (A-B) Density plots before and after the removal of one control sample with a distinct beta density distribution. (C-D) Heatmap plots of the singular value decomposition analysis are presented for before (C) and after (D) the removal of batch effects. Color represents significance level. A darker color indicates a stronger association between a variable (row name) and a principal component (column name). All batch variables(array, slide and sample plate effects) become uncorrelated after batch correction.

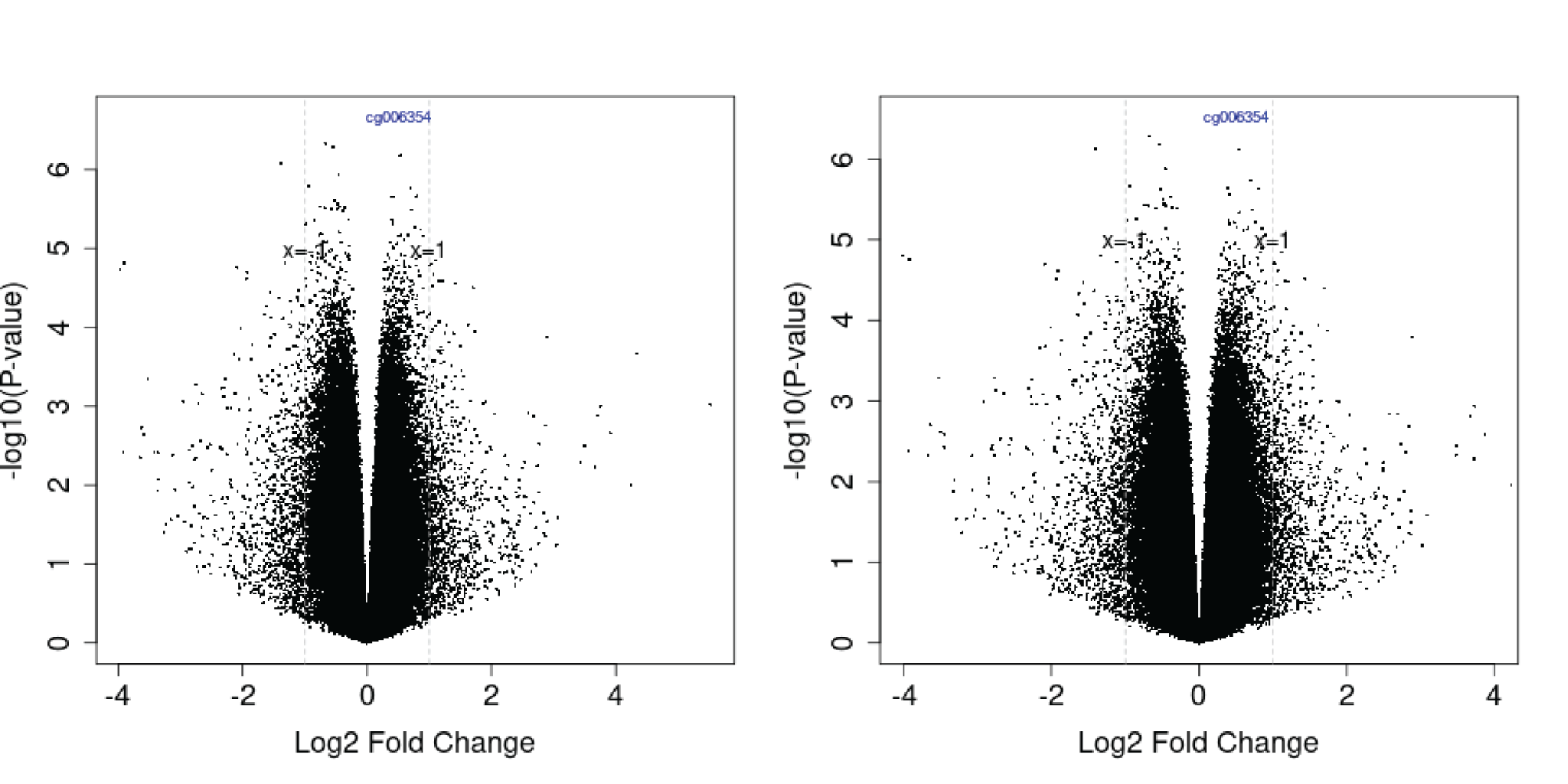

**Supplementary Figure 3**: **DNA methylation result of in-house data before (left)and after(right) adjustment of smoking**. This did not change the observation of a lack of significant CpGs from the cord blood associated with severe PE.

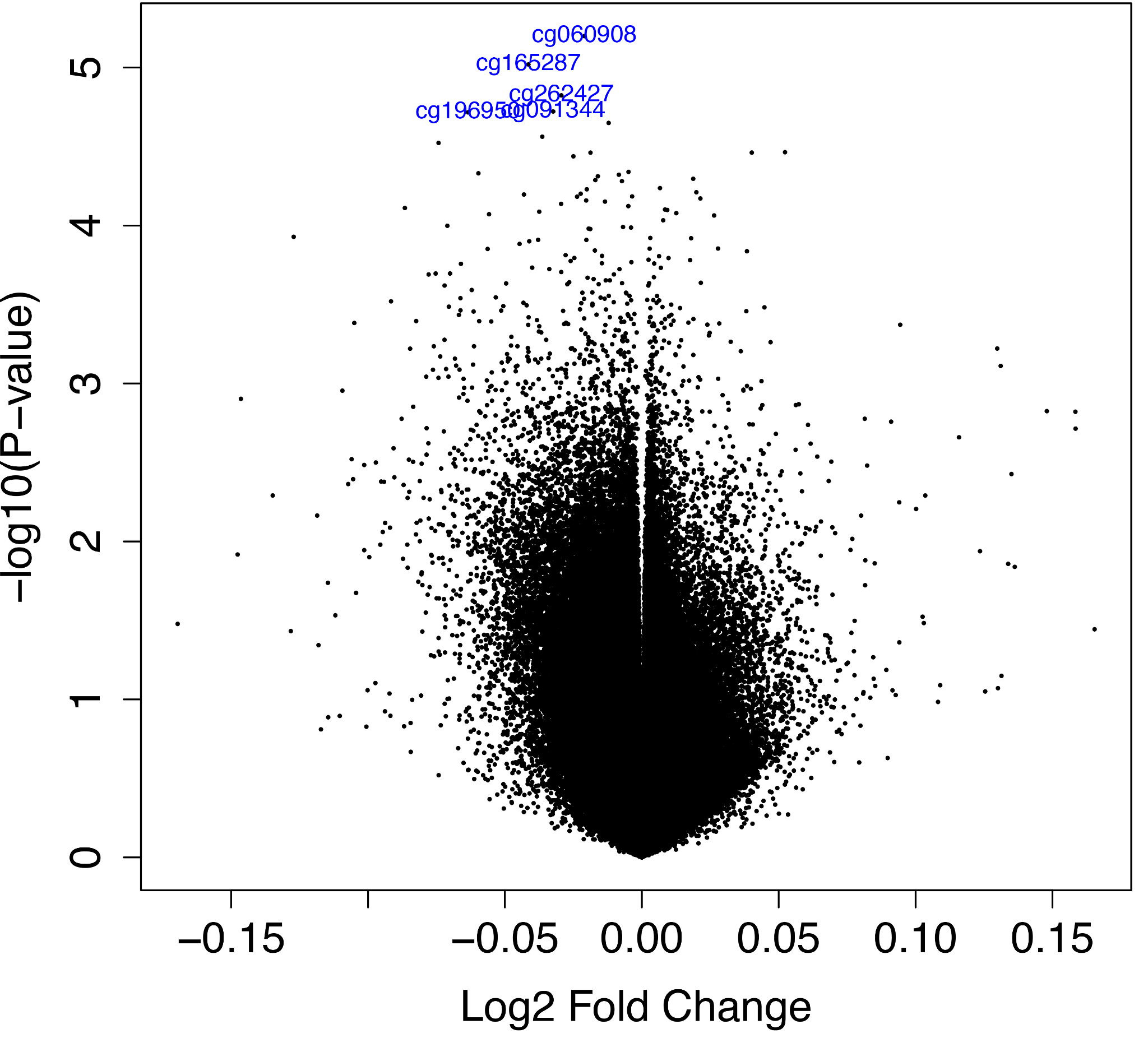

**Supplementary Figure 4: Differential methylation results without confounding adjustment for the study of Kashima K et al.** No significant CpGs were found before any confounding adjustment.

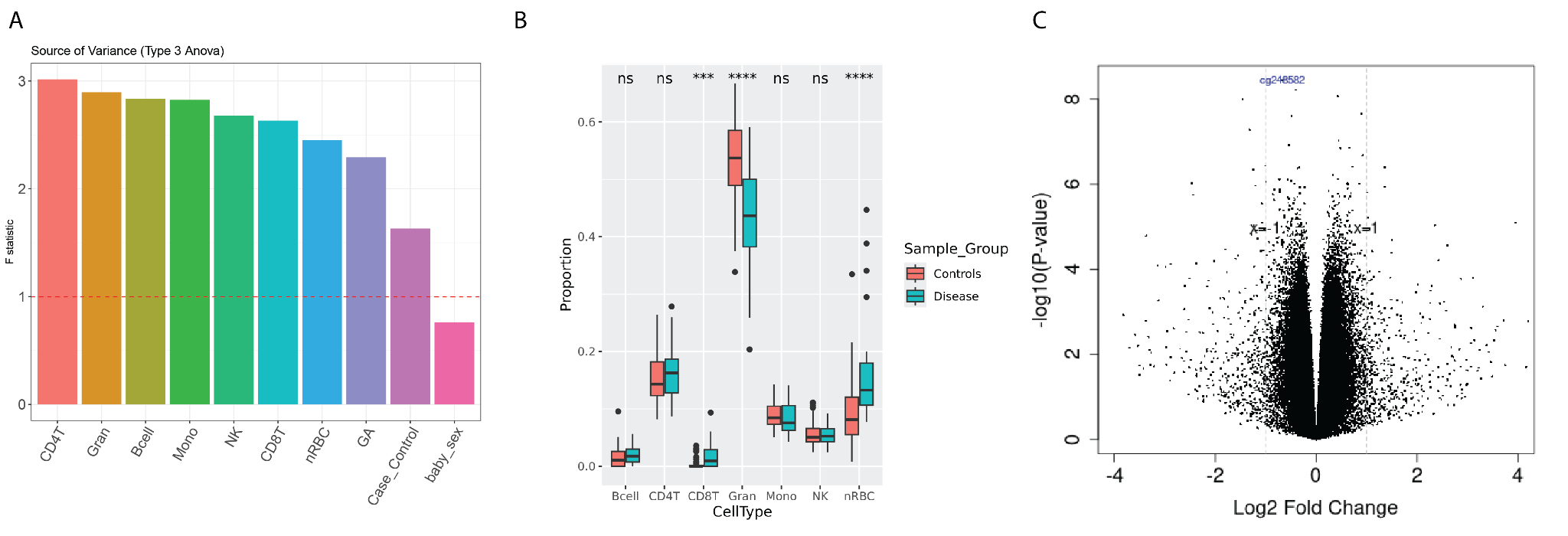

**Supplementary Figure 5: Differential methylation result after adding data from Fernando et al., (**[**GSE66459**](https://www.ncbi.nlm.nih.gov/geo/query/acc.cgi?acc=GSE66459)**).** Fernando et al. contains 11 idiopathic preterm samples and 11 full-term samples(see Methods). A) The Source of Variance (SOV) analyses were conducted on cell types and clinical covariates. B) Estimated cell type proportion in the merged dataset. C) volcano plot of differential methylation result adjusted for clinical confounders and estimated cell proportion.

**Supplementary Tables**

**Supplementary Table 1: Multiple linear regression of each cell type on clinical variables**

| **Supplementary Table 1: Linear regression of each cell type on clinical variables** | | | | | | | | | | | | | | |
| --- | --- | --- | --- | --- | --- | --- | --- | --- | --- | --- | --- | --- | --- | --- |
|  | *CD8T* | | *CD4T* | | *B cell* | | *Granulocyte* | | *Monocyte* | | *Natural Killer* | | *nRBC* | |
|  | *Coefficient* | *P-value* | *Coefficient* | *P-value* | *Coefficient* | *P-value* | *Coefficient* | *P-value* | *Coefficient* | *P-value* | *Coefficient* | *P-value* | *Coefficient* | *P-value* |
| ***(Intercept)*** | 6.87E-02 | 0.279 | 2.89E-01 | 0.038 | 9.89E-02 | 0.008 | -3.27E-01 | 0.118 | -1.54E-01 | 0.052 | 1.35E-01 | 0.022 | 9.38E-01 | 0.000 |
| ***PE*** | 1.81E-02 | **0.017** | 7.76E-03 | 0.627 | -2.20E-04 | 0.959 | -4.66E-02 | 0.059 | 1.36E-02 | 0.140 | -6.27E-03 | 0.355 | 9.06E-03 | 0.672 |
| ***Age*** | 8.64E-06 | 0.987 | 4.00E-04 | 0.728 | 1.14E-05 | 0.970 | 1.40E-03 | 0.423 | 7.85E-04 | 0.234 | -4.36E-04 | 0.372 | -2.39E-03 | 0.125 |
| ***GA*** | -1.12E-03 | 0.444 | -2.74E-03 | 0.387 | -2.20E-03 | **0.011** | 2.16E-02 | **0.000** | 5.72E-03 | **0.002** | -2.12E-03 | 0.118 | -2.20E-02 | **0.000** |
| ***BMI*** | -6.55E-04 | 0.064 | -1.27E-03 | 0.096 | -2.73E-04 | 0.178 | -2.90E-04 | 0.799 | 2.74E-04 | 0.523 | -2.78E-04 | 0.384 | 2.51E-03 | **0.015** |
| ***Caucasian*** | 7.87E-04 | 0.916 | 1.12E-02 | 0.487 | 5.57E-03 | 0.197 | 1.29E-03 | 0.958 | -1.14E-02 | 0.218 | 4.58E-03 | 0.501 | -1.42E-02 | 0.510 |
| ***Pacific Islander*** | 8.47E-03 | 0.254 | 2.00E-02 | 0.213 | 9.26E-03 | **0.033** | 2.13E-02 | 0.382 | -1.47E-03 | 0.871 | -3.31E-03 | 0.624 | -5.78E-02 | **0.009** |
| ***Parity*** | 1.06E-04 | 0.954 | 5.36E-03 | 0.179 | 5.43E-04 | 0.607 | -3.63E-03 | 0.546 | 2.11E-03 | 0.352 | -4.13E-04 | 0.805 | -3.30E-03 | 0.533 |

**Supplementary Table 2: Top 20 Differentially Methylated Regions**

|  | seqnames | start | end | width | strand | value | area | cluster | indexStart | indexEnd | L | clusterL | p.value | fwer | p.valueArea | fwerArea |
| --- | --- | --- | --- | --- | --- | --- | --- | --- | --- | --- | --- | --- | --- | --- | --- | --- |
| DMR_2 | chr5 | 135,000,000.00 | 135,000,000.00 | 920 | * | 1.23391 | 17.27474 | 379040 | 97921 | 97934 | 14 | 14 | 2.36E-06 | 0.04 | 6.13E-06 | 0.10 |
| DMR_1 | chr21 | 47,604,052.00 | 47,605,174.00 | 1122 | * | -1.45296 | 13.07662 | 299226 | 79019 | 79027 | 9 | 9 | 9.43E-07 | 0.02 | 1.44E-05 | 0.22 |
| DMR_8 | chr17 | 5,402,883.00 | 5,403,907.00 | 1024 | * | -0.80212 | 7.219061 | 191023 | 49988 | 49996 | 9 | 14 | 3.39E-05 | 0.44 | 1.07E-04 | 0.81 |
| DMR_3 | chr17 | 48,585,216.00 | 48,585,575.00 | 359 | * | 1.062775 | 6.376653 | 202519 | 54250 | 54255 | 6 | 13 | 1.04E-05 | 0.16 | 1.60E-04 | 0.89 |
| DMR_12 | chr1 | 154,000,000.00 | 154,000,000.00 | 669 | * | 0.446431 | 6.250031 | 28477 | 8094 | 8107 | 14 | 15 | 6.86E-05 | 0.74 | 1.70E-04 | 0.90 |
| DMR_13 | chr8 | 11,666,281.00 | 11,666,810.00 | 529 | * | -0.64207 | 5.778594 | 445420 | 118721 | 118729 | 9 | 9 | 9.36E-05 | 0.74 | 2.20E-04 | 0.94 |
| DMR_14 | chr6 | 28,911,464.00 | 28,912,166.00 | 702 | * | -0.56824 | 5.68238 | 394630 | 102270 | 102279 | 10 | 12 | 9.10E-05 | 0.75 | 2.31E-04 | 0.95 |
| DMR_16 | chr8 | 22,132,678.00 | 22,133,076.00 | 398 | * | 0.665114 | 5.320914 | 446968 | 119003 | 119010 | 8 | 13 | 0.000111 | 0.78 | 2.80E-04 | 0.96 |
| DMR_9 | chr22 | 51,016,386.00 | 51,017,166.00 | 780 | * | -0.31714 | 5.074293 | 310000 | 81738 | 81753 | 16 | 18 | 3.39E-05 | 0.45 | 3.20E-04 | 0.98 |
| DMR_19 | chr4 | 81,118,188.00 | 81,118,794.00 | 606 | * | -0.64186 | 5.134847 | 350421 | 91322 | 91329 | 8 | 8 | 0.000128 | 0.83 | 3.09E-04 | 0.98 |
| DMR_21 | chr5 | 179,000,000.00 | 179,000,000.00 | 615 | * | -0.39445 | 5.127847 | 387844 | 100451 | 100463 | 13 | 14 | 0.000103 | 0.87 | 3.09E-04 | 0.98 |
| DMR_5 | chr6 | 29,911,036.00 | 29,911,104.00 | 68 | * | -0.96481 | 4.824052 | 394963 | 102826 | 102830 | 5 | 20 | 2.38E-05 | 0.34 | 3.71E-04 | 0.98 |
| DMR_22 | chr7 | 142,000,000.00 | 142,000,000.00 | 344 | * | -0.6399 | 4.479273 | 438856 | 117654 | 117660 | 7 | 17 | 0.000159 | 0.89 | 4.59E-04 | 0.99 |
| DMR_28 | chr17 | 33,759,929.00 | 33,760,419.00 | 490 | * | -0.4435 | 4.434984 | 197327 | 52091 | 52100 | 10 | 17 | 0.000187 | 0.93 | 4.70E-04 | 0.99 |
| DMR_30 | chr6 | 26,225,246.00 | 26,225,767.00 | 521 | * | -0.43291 | 4.329146 | 393846 | 101472 | 101481 | 10 | 13 | 0.000196 | 0.94 | 5.04E-04 | 0.99 |
| DMR_34 | chr14 | 24,779,959.00 | 24,780,734.00 | 775 | * | 0.38551 | 4.24061 | 137001 | 35021 | 35031 | 11 | 15 | 0.000178 | 0.95 | 5.34E-04 | 0.99 |
| DMR_35 | chr1 | 206,000,000.00 | 206,000,000.00 | 536 | * | -0.40438 | 4.043765 | 38481 | 10682 | 10691 | 10 | 14 | 0.000224 | 0.96 | 6.09E-04 | 0.99 |
| DMR_38 | chr2 | 30,669,385.00 | 30,669,952.00 | 567 | * | 0.550183 | 3.851279 | 246773 | 67490 | 67496 | 7 | 11 | 0.000304 | 0.97 | 6.81E-04 | 1.00 |
| DMR_39 | chr1 | 110,000,000.00 | 110,000,000.00 | 251 | * | -0.47774 | 3.821909 | 24488 | 6686 | 6693 | 8 | 10 | 0.000369 | 0.97 | 6.94E-04 | 1.00 |
| DMR_40 | chr15 | 69,744,390.00 | 69,744,850.00 | 460 | * | -0.53819 | 3.767325 | 161393 | 41643 | 41649 | 7 | 12 | 0.000329 | 0.98 | 7.21E-04 | 1.00 |
